# Patient and Provider Perceptions of a Community-Based Accompaniment Intervention for Adolescents Transitioning to Adult HIV Care in Urban Peru: A Qualitative Analysis

**DOI:** 10.1101/2022.04.11.22273102

**Authors:** Jerome T. Galea, Milagros Wong, Brennan Ninesling, Alicia Ramos, Liz Senador, Hugo Sanchez, Lenka Kolevic, Eduardo Matos, Eduardo Sanchez, Renato A. Errea, Andrew Lindeborg, Carlos Benites, Leonid Lecca, Sonya Shin, Molly F. Franke

## Abstract

**Introduction:** Adolescents living with HIV (ALWH) experience higher mortality rates compared to other age groups, exacerbated by suboptimal transition from pediatric to adult HIV care in which decreased adherence to antiretroviral treatment (ART) and unsuppressed viremia are frequent. Care transition—a process lasting months or years—ideally prepares ALWH for adult care and can be improved by interventions that are youth-friendly and address psychosocial issues affecting ART adherence; however, such interventions are infrequently operationalized. Community-based accompaniment (CBA), in which laypeople provide individualized support and health system navigation, can improve health outcomes among adults with HIV. Here, we describe patient and provider perceptions of a novel HIV CBA intervention called “PASEO” for ALWH in Lima, Peru.

**Methods:** PASEO consisted of six core elements designed to support ALWH before, during, and after transition to adult HIV care. Community-based health workers provided tailored accompaniment for ALWH aged 15-21 years over 9 months, after which adolescent participants were invited to provide feedback in a focus group or in-depth interview. HIV care personnel were also interviewed to understand their perspectives on PASEO. A semi-structured interview guide probing known acceptability constructs was used. Qualitative data were analyzed using a Framework Analysis approach and emergent themes were summarized with illustrative quotes.

**Results:** We conducted 5 focus groups and 11 in-depth interviews among N=26 ALWH and 9 key-informant interviews with HIV care personnel. ALWH participants included those with both vertically- and behaviorally acquired HIV. ALWH praised PASEO, attributing increased ART adherence to the project. Improved mental health, independence, self-acceptance, and knowledge on how to manage their HIV were frequently cited. HIV professionals similarly voiced strong support of PASEO. Both ALWH and HIV professionals expressed hope that PASEO would be scaled. HIV professionals voiced concerns regarding financing PASEO in the future.

**Conclusion:** A multicomponent CBA intervention to increase ART adherence among ALWH in Peru was highly acceptable by ALWH and HIV program personnel. Future research should determine the efficacy and economic impact of the intervention.

## Introduction

HIV is a manageable disease from which mortality should be rare; however, adolescents aged 10-19 living with HIV (ALWH)—about 1.8 million globally [1]—experience worse health outcomes than other age groups. HIV is the second leading cause of death among adolescents aged 10-19 years [1-3], and among youth aged 15-19 years, HIV-associated mortality is rising [4]. For ALWH, becoming an adult is an especially precarious period. This is because, in addition to the biopsychosocial changes during adolescence, ALWH also transition from pediatric to adult HIV care and are frequently “lost in transition,” especially in low-resource settings [5].

Care transition is a process lasting months or years, that ideally ensures that ALWH are virally suppressed before, during, and after transition [6]. Achieving viral suppression pre-transition is vital; a 2020 systematic review (N=24 studies) of care transition research among ALWH found the worst health outcomes among those with unsuppressed viremia [7]. However, in ALWH, viral suppression rates are disappointing. For example, among a cohort of 1411 U.S. adolescents aged 13-29 years with mostly behaviorally acquired HIV, after a median follow up of 5 months, only 34% were retained in HIV care and started antiretroviral therapy (ART), and just 12% achieved viral suppression [8]. Another study among N=401 Spanish adolescents with vertically acquired HIV found that 3.5% (n=14) had died within ten years post-transition [9].

Knowledge on the factors affecting care transition among ALHW is rapidly expanding [10-16], though data are primarily from Africa, the U.S., and Europe. Nonetheless, the need for interventions that are individualized, youth-friendly, collaborative between youth and medical providers, and responsive to ALWH’s mental/psychosocial needs are consistently noted [17-22]. Several studies highlight ALWH’s transition experiences, emphasizing attention to issues related to becoming an adult (e.g., learning core competencies needed to manage HIV autonomously [23]), and not limited to only ART adherence [24]. Only one review included Latin America [25], but again found that structural- and individual-level barriers (e.g., lack of family support, HIV stigma, poverty, later HIV disclosure), negatively affect ALWH’s treatment outcomes.

Despite the evidence on facilitators of successful care transition, research to improve ALWH’s experiences is scant. An approach that enhances HIV outcomes among adults—community-based accompaniment (CBA)—holds promise [26]. CBA trains lay people to provide support, education, and assistance navigating the HIV care system. For example, adults living with HIV in Peru and Rwanda who received HIV CBA had higher suppressed viremia rates and improved mental and social health outcomes relative to control groups [27-30]; however, CBA for ALWH transitioning to adult HIV care has not yet been tested.

In Peru, approximately 76,000 people live with HIV, of which 11,000 are aged 15-24 years [31]. ALWH in Peru are diverse, consisting of youth with perinatally and behaviorally acquired HIV, young men who have sex with men (MSM), and transgender women; the epidemic is concentrated among the latter two groups, with those aged 19-26 years most affected [31]. Here, we describe ALWH’s and HIV provider perspectives of a novel CBA intervention for ALWH transition to adult HIV care in Lima, Peru.

## Methods

### Participants and procedures

During 2019-2021, we pilot-tested a CBA intervention called “PASEO” (‘crossing’ or ‘passage’ in Spanish) implemented by the community-based organization Socios En Salud (SES) in Lima. Building on barriers to ART adherence experienced by Peruvian ALWH [32], PASEO provided CBA to ALWH aged 15-22 years commencing or already taking ART as they prepared for and/or transitioned to adult HIV care. Adolescents that had already transitioned to but disengaged from adult HIV care also participated.

The PASEO intervention is described elsewhere [33]. Briefly, PASEO consisted of six core components delivered during a six-month intensive phase followed by a three-month step-down phase. The components were: 1. Health system navigation and clinic visit accompaniment; 2. Social support groups; 3. Screening and referral to mental health services; 4. Resolution of acute needs (e.g., psychosocial, medical, housing, transportation); 5. Health education and skills-building; and 6. Individualized adherence support, including Directly Observed Treatment (DOT), as needed.

ALWH with both vertically- and behaviorally acquired participated, including cisgender males and females, males identifying as homosexual, and transgender females. Some in-person PASEO components were delivered virtually when the COVID-19 pandemic emerged in March 2020, at which point participants had received between 1.4-5.3 months of the intervention [34].

An ALWH-comprised Youth Advisory Board gave feedback/suggestions throughout the study. Ethics Committees of the following institutions gave ethical approval for this work: Hospital Nacional Arzobispo Loayza, Lima, Peru [approval number: 11698]; Instituto Nacional de Salud del Niño, Lima, Peru [approval number: 00213]; Hospital Nacional Hipólito Unanue, Lima, Peru [approval number: 17845]; Harvard Medical School, Boston, United States [approval number: 19-0086]. Written informed consent, including permission to publish study results stripped of personally identifying information, was obtained from adolescents ≥18 years of age and from guardian(s) of adolescents <18 years with informed assent. Consent was waived for adolescents aged <18 years without guardians; they provided informed assent. PASEO was registered with ClinicalTrials.gov (Identifier: NCT05022706).

### Data collection

Thirty ALWH participated in PASEO, and all were invited to participate in a focus group or in-depth interview at study completion, conducted by MW, the PASEO study coordinator, and a registered nurse, trained in qualitative research methods. Focus group composition was determined *a priori*, based on participants’ shared characteristics (Table 1). In-depth interviews were also conducted among participants for a deeper understanding of their experience. Additionally, we conducted key informant interviews with HIV care personnel (physicians, peer counselors, psychologists, program officials).

**Table 1:** Focus groups and in-depth interview composition among adolescent participants in PASEO.

| Participant type and/or experience during participation in PASEO |
| --- |
| A. Adolescents that regularly participated in **all** PASEO intervention components |
| B. Adolescents that did **not** regularly participate in all PASEO intervention components |
| C. Adolescents that were pregnant during PASEO |
| D. Adolescents that transitioned from pediatric to adult care during PASEO |
| E. Adolescents that initiated HIV antiretroviral treatment just prior to PASEO |

The focus groups, in-depth, and key-informant interviews followed semi-structured interview guides exploring eight feasibility areas [35] (Table 2). Data collection occurred via video conferencing due to COVID-19; for 3 participants, interviews occurred via encrypted text messages because they relocated outside Lima and lacked internet. Interviews and focus groups lasted approximately 60-minutes, were recorded, transcribed verbatim, and analyzed using Dedoose [36].

**Table 2:**
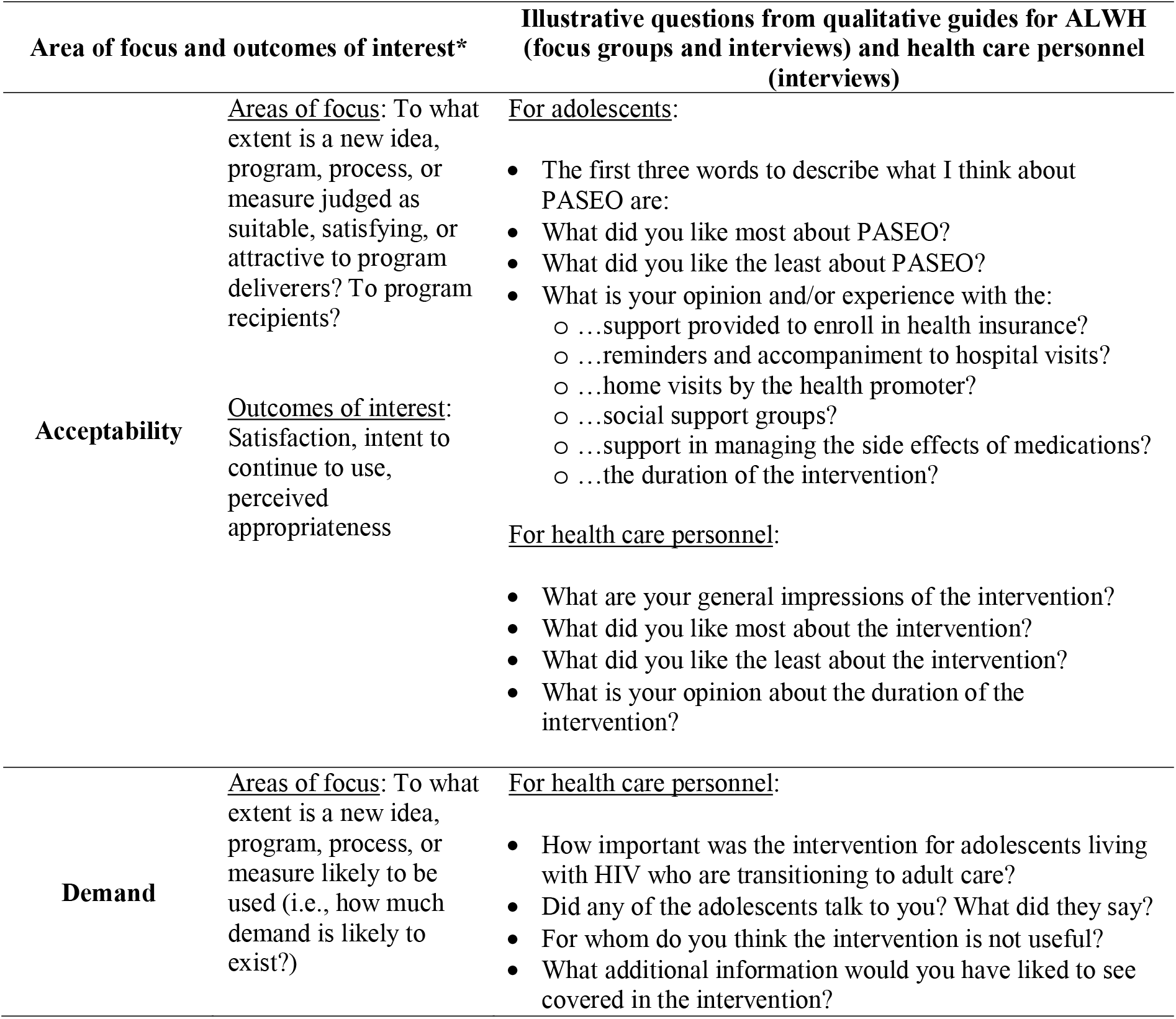
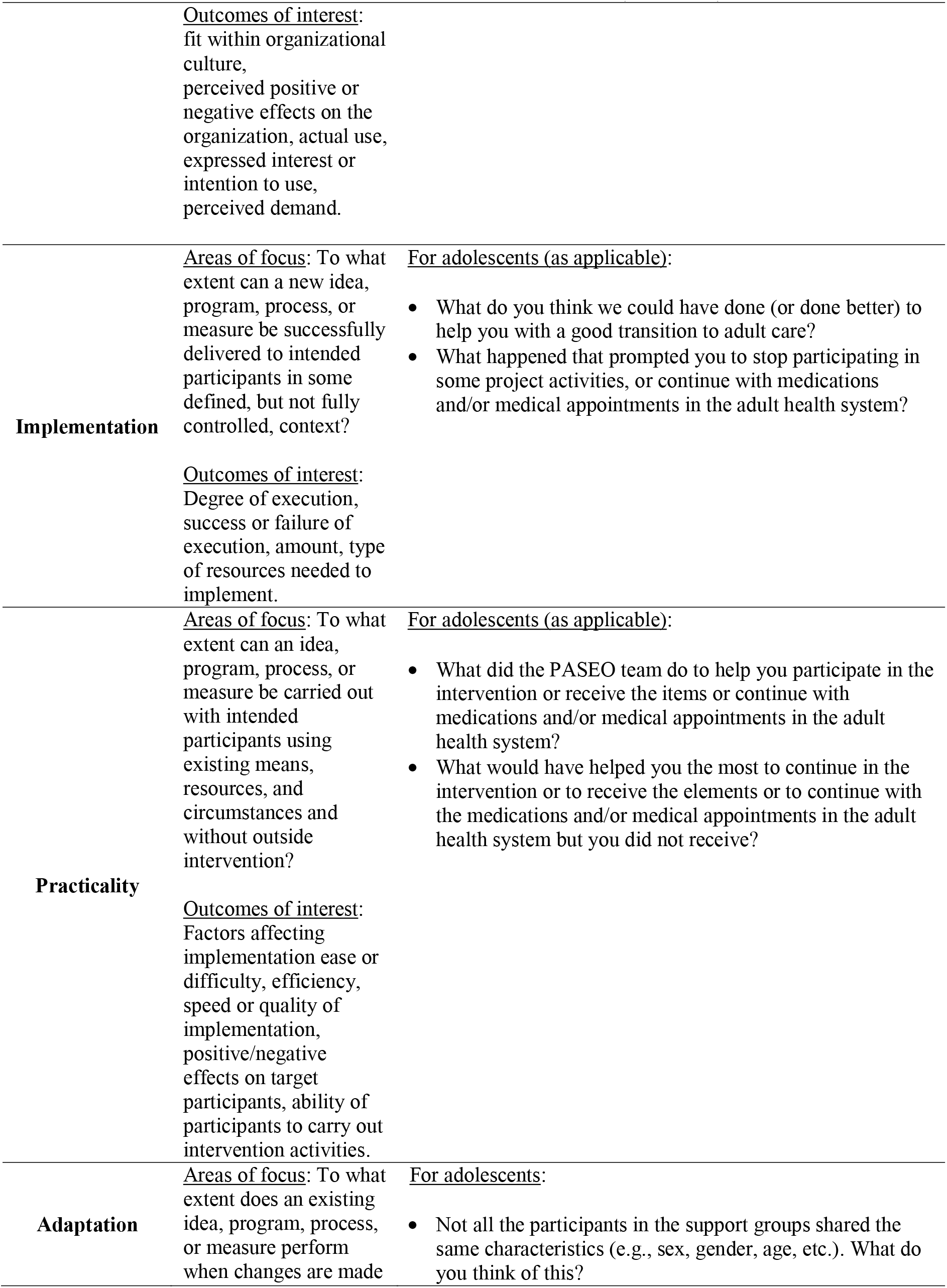
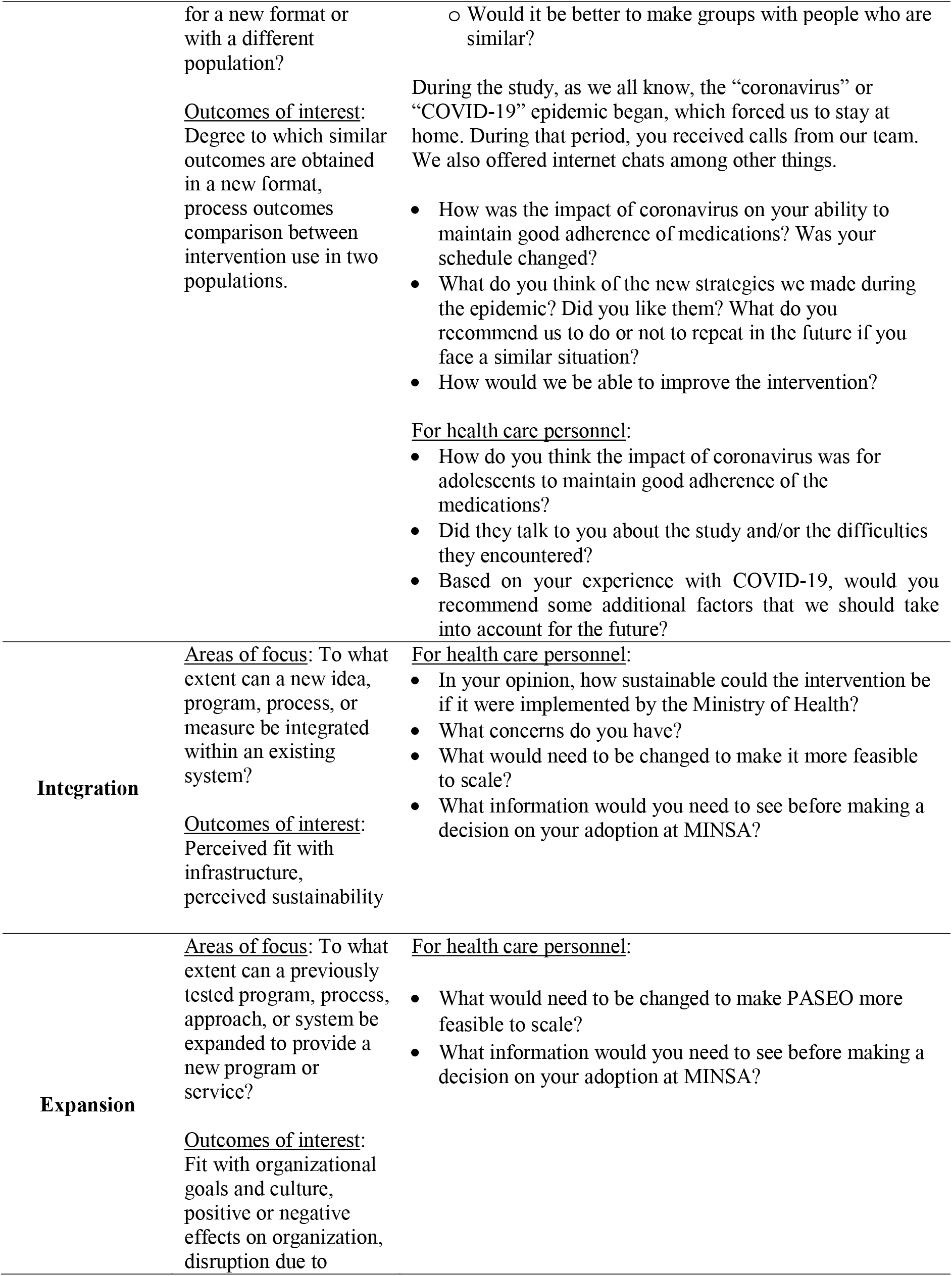
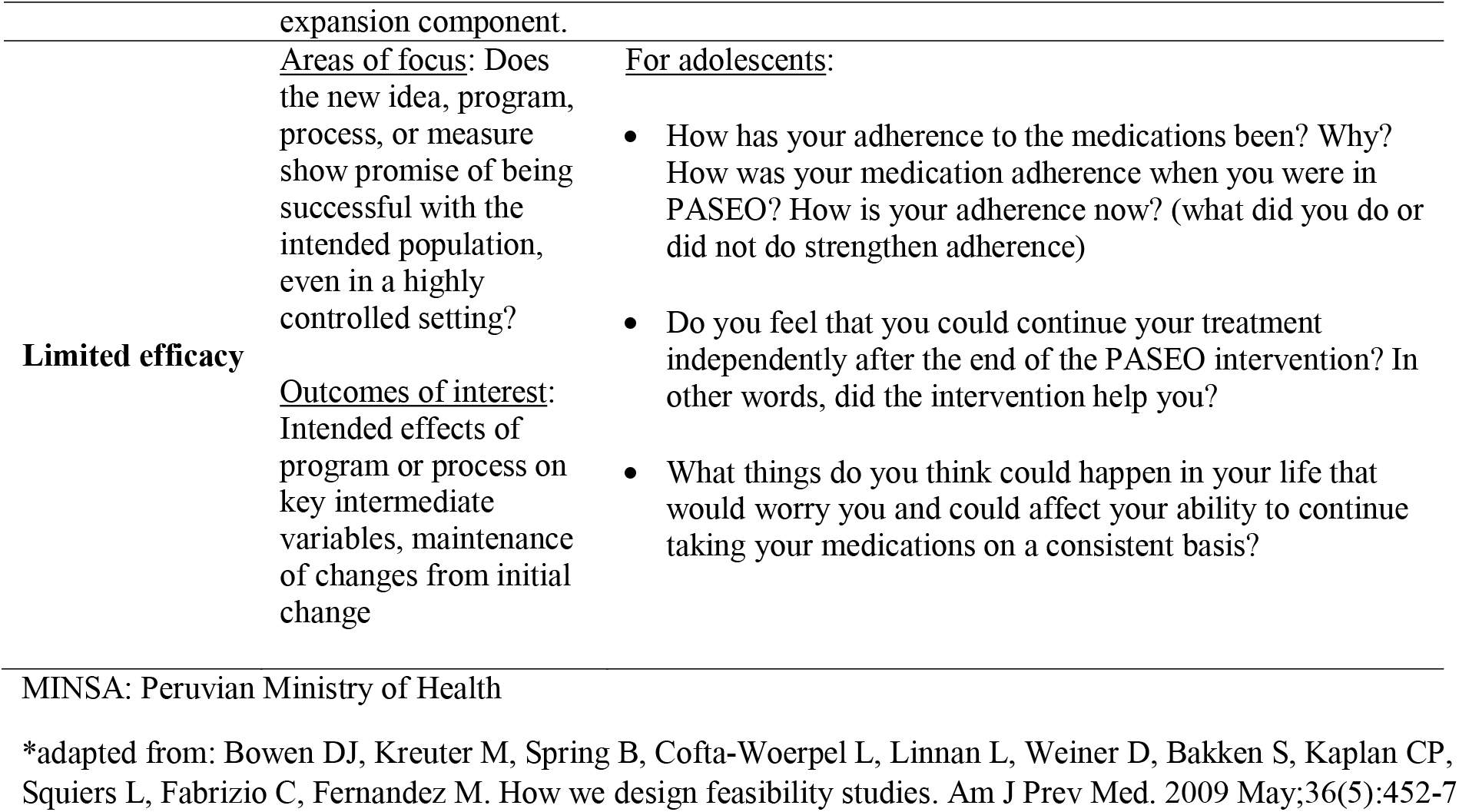
Areas of focus, outcomes of interest, and illustrative questions used in qualitative data collection guides used in focus groups and interviews to assess the acceptability of the PASEO intervention*.

### Data Analysis

The qualitative analysis team was comprised of two coders (MW and BN) trained in framework analysis [37]. Analysis began with the coders independently reading and coding five transcripts using a preliminary codebook derived from the interview guides, adding *de novo* codes as needed. Separate codebooks were created for ALWH and HIV personnel. Next, the team compared the five coded transcripts and harmonized coding by consensus. Reports were generated for each code, and related text across all transcripts was extracted into matrices for granular analysis. Finally, crosscutting themes were identified and reported using illustrative quotes. The Consolidated Criteria for Reporting Qualitative Data Checklist (COREQ) was completed to enhance rigor and transparency (Additional File).

## Results

### Participant characteristics

Twenty-six ALWH participated in a focus group or in-depth interview (n=15 and n=11, respectively); n=4 participants were lost to follow-up/non-responsive to invitations to participate. Fifteen (58%) participants were male of which nearly all (93%) identified as cisgender. Most (69%) ALWH acquired HIV during early childhood and over half (62%) had a parent die from HIV (Table 3).

**Table 3:** Characteristics of PASEO ALWH that participated in a focus group or in-depth interview (N=26)

| Characteristics | N (%) |
| --- | --- |
| **Sex assigned at birth** | Male 15 (58) |
|  | Female 11 (42) |
| **Sexual orientation** | Homosexual 9 (35) |
|  | Heterosexual 17 (65) |
| **Gender** | Cisgender male 14 (54) |
|  | Cisgender female 11 (42) |
| **Timing of HIV acquisition** | Early childhood 18 (69) |
|  | Recent 8 (31) |
| **Death of parent** | Both parents 11 (42) |
|  | One parent 5 (19) |
| **Currently living with/in** | Any family member 6 (23) |
|  | Father and/or mother 8 (31) |
|  | Group home 5 (19) |
|  | Partner or friend 7 (27) |
| **Lifetime history of residence in group home for children with HIV/AIDS** | 7 (27) |
| **Reason for referral to PASEO** | Transitioning from pediatric to adult HIV care 6 (23) |
|  | Disengaged from HIV care or poor ART adherence after care transition 12 (46) |
|  | Initiating ART 8 (31) |

ART: antiretroviral medications

Nine key-informant interviews were conducted with HIV program personnel: three physicians, one nurse, two peer counselors, two psychologists, and a group home coordinator.

### Qualitative findings

For ALWH, qualitative data are organized under two domains, <u>Domain 1</u>: Overall acceptability and <u>Domain 2</u>: ALWH’s experiences with PASEO’s six core components. For HIV personnel, data are grouped under <u>Domain 1</u>: Opinions of PASEO and <u>Domain 2</u>: Post-PASEO recommendations. Each category, theme, and sub-theme (as applicable) is presented with illustrative quotes. For adolescent data, the attributions following the quote include focus group or in-depth interview (FG or IDI) and participant experience type (per Table 1, A, B, C, D, E). For HIV professionals’ data, due to their small number, quote attributions do not include job role to preserve anonymity.

### Adolescents

#### Domain 1, Experiences during PASEO - Overall acceptability

##### Likes

Overall, participants were positive about PASEO, repeatedly reporting feeling that PASEO was effective, and built camaraderie among ALWH. One participant stated, “If I had not [enrolled in PASEO], I would still be without treatment; of that, I am sure” (FG, A). Another participant summed up their experience in three words: “When I think of PASEO, the three words that come to mind are]: community, teaching, help. Teaching because they taught us many things, things that I didn’t know. Community because the program connected us with other people just like us” (FG, B).

ALWH emphasized the social support aspect of PASEO; this participant found that PASEO had arrived at just the right time in their life:

> …to be honest, I could say that I was totally lost—disoriented—and I didn’t know how I was going to do it. Sometimes, I got so low that I felt horrible, but knowing that I had someone who supported me, that helped me, emotionally more than anything, that was what I needed most at that time. (FG, E)

PASEO also had the impact of reducing social isolation, wherein new acquaintances were nonjudgmental, and anything could be discussed without fear:

> …before [PASEO] I just stayed in my living room alone. But then I met new people and felt free to talk about things and not be judged, like any topic. They didn’t judge you when you said something, whether it was right or wrong.” (IDI, C)

##### Dislikes

Dislikes were logistical in nature, relating to the time required to participate in PASEO or the pivot to virtual activities when COVID-19 emerged. One participant found difficulty with the group sessions due to academic commitments, stating, “What I didn’t like about PASEO was that the sessions didn’t adapt very well to my schedule that I had to have for my virtual classes” (IDI, E). Another participant disliked the virtual sessions due to COVID-19: “The only thing I didn’t like was the pandemic, because due to the pandemic we couldn’t meet in person, and everything was virtual. […] I liked [PASEO] before the pandemic a thousand times more” (FG, C).

Another issue with virtual activities, necessary due to COVID, was related to technical problems such as limited access to Wi-Fi or cellular data. One participant voiced, “Well, [I didn’t like] participating with video because sometimes my phone’s megabytes would run out quickly, and then I couldn’t use it. After my data ran out, I had to use my neighbor’s Wi-Fi when I wanted to […] get into the [online support group]” (IDI, B).

##### Intervention duration

When participants were asked their opinion regarding the duration of PASEO, a consistently stated preference for a longer intervention period emerged, though some participants voiced support for a variable length intervention responsive to the needs of the adolescent. In this example, the ALWH used their own experience of sub-optimal ART adherence to make the case for individualized duration of support:

> …I didn’t take the pills, or if I took them, I took them every other day; I forgot at night or in the morning because I had other things to do. I thought about other things and that’s why, right? […] I don’t know, I guess like [another participant] said, [the duration] depends on the person that needs the support. (FG, D)

Recommendations specifically for a longer intervention duration were often framed in the context of having more time to form/maintain social bonds with other PASEO peers, and some participants expressed sadness that PASEO had ended:

> …thinking about being close to finishing the project really affects me a lot, because I love the meetings that we have. I listen to everyone and compare myself to each person, and I realize that I’m not the only person who has suffered or had those thoughts, and I like that. (FG, A)

Another participant expressed, “…the end of the project is painful, and it’s a little sad. But the truth is that the times that we’ve had have been really, really good” (FG, A).

#### Domain 2: Experiences with the 6 PASEO core components

##### Component 1 - Health system navigation and clinic visit accompaniment

Participants praised the support received from the health promoter (“*promotora*” in Spanish), with three interrelated themes emerging: immediate assistance and training; attenuating prior bad experiences with the health system; and ongoing, trust-based working relationships. This participant notes how she learned to make clinic visit appointments on her own:

[My promotora] was like a guide. For example, with the [health insurance program] she told me, ‘Look, you have to be there early, you have to be persistent, and explain your situation well.’ When it was time to make appointments […] she made me do it so that I would know what to do. Then if they didn’t give me the appointment or if I couldn’t make the payment, she would show up and fight with who she needed to get the appointment. (FG, A)

For another participant, accompaniment appeared to buffer against previous bad experiences with the health system, restoring respect by health personnel: “… [in the hospital where I receive care] everyone treated me really poorly the several times that I went alone. I recently went for a family planning visit, and the doctor treated me as if I was from another world. But when you have company, they respect you a little more” (FG, B). Feedback from one adolescent echoed the common view by ALWH that the community health workers were more than project staff; they were trusted friends who stood by their side, saying, “I am very grateful because [my promotora] was there with me even though I refused to go back to my treatment. She was there with me until I became undetectable, and I owe it all to her because she was there not only as a worker, but as my friend” (FG, A).

##### Component 2 - Social support groups

Participants spoke profusely of the value of PASEO’s support groups; however, there were disparate views on group composition. Some participants felt that groups comprised of all ALWH (regardless of sex, gender, and HIV acquisition route) was a strength because they could learn from others with similar but different experiences: “…I learned a lot, like at least one thing I feared was not being able to have a baby, and there I learned that I can have one, because there were several guys, several girls [diagnosed with HIV] that had a baby” (IDI, E).

Participant: “I liked how [the groups] were, I liked the group that I was in. We had great conversations, and we became friends.”

Interviewer: “So [you liked the] varied groups, with men, women, vertical transmission, or others.”

Participant: “We’re all the same.” (FG, B)

In contrast, some participants felt that grouping ALWH with similar experiences could be beneficial, for example, those just starting HIV treatment:

> …for me, it would have been a little better if it had been the same people, those like me, to see that they were going through the same thing. Those that had just started treatment and things like that. Then, there’d be more things we have in common because, I don’t know, you get to help them faster and they can help themselves too. (FG, E)

##### Component 3: Screening and referral to mental health services

ALWH unanimously praised the PASEO’s emphasis on mental wellbeing, often framing their experiences in the context of personal growth and acceptance of living with HIV—even as a transformative experience—as in this example:

[The psychologist] asked me, ‘Have you really accepted yourself as you are, have you really accepted yourself?’ And that’s when it struck me, I felt bad at that time, and I left the question there in the air, meditating on it for a while, realizing what the problem was. That was the problem, no? That I didn’t really accept my diagnosis, I hadn’t come to terms with the fact that I was an HIV carrier, that I have had to live this life. (IDI, A)

Another participant spoke of a traumatic past of being bullied, and how PASEO helped them develop self-acceptance: “I grew up getting bullied a lot, you know, they bullied me a lot and then made me angry at what I have, until I got here and learned many things from the people here and accepted what I have” (FG, B).

##### Component 4 - Resolution of acute needs

Participants spoke positively about the support they received, again echoing a special connection with their *promotora*, and their ability to assess and address adolescents’ acute needs that impacted ART adherence, as for this participant who was homeless:

…I was going to die because I was on the street, and I was going to [northern Peru]. […] [The PASEO team] took me to the hospital and that’s where I also learned about the project. They saved my life because at that point I wasn’t thinking about starting my medication again. (FG, A)

##### Component 5 - Health education and skills-building

For this core element, participants’ experiences overlapped with thematic content for mental health and social support as they related to personal growth. In the following example, a pregnant ALWH spoke about learning how to manage her specific health needs, and with Component 1, also practiced self-directing her care:

…when I was pregnant and had to go to hematology, I didn’t know much about what that was for. [My promotora] sometimes asked the doctor questions, and the doctor told her about my hemoglobin, about what I must take, and those things. […] I usually kept quiet but ever since I saw [my promotora] ask the doctor questions and things like that, about her doubts, well, now I sometimes ask the doctor questions too. I learned that from her, how to speak to the doctor, because I was practically always mute in the hospital. (FG, C)

##### Component 6 – Individualized adherence support with directly observed treatment (DOT)

For participants that required additional adherence support, and received DOT, their experiences were characterized as both a reminder to take ART and as an exercise in learning personal responsibility for ART adherence:

> …in my case, sometimes I didn’t take my medication because I forgot to, or I lost track of time. So then with the lady [from PASEO] ‘s calls on WhatsApp, I knew that at 8:00 PM I had to take my pill, from my first call and on. So now, like all the other girls and guys here, I think it’s one’s own responsibility. (FG, D)

In the following example, DOT—and the experience of being confronted by the CHW for non-adherence—was even framed as a reminder and also an act of support:

> … I was very forgetful, I forgot to take my pills, I forgot my appointments, I forgot everything. […] More than anyone, the lady [from PASEO] helped me because she yelled at me every time I forgot, well she chided me. She would ask me why I forgot, and she chided me every time I forgot. I felt that she cared about me more than anyone else, more than my family. (FG, B)

### HIV Personnel

#### Domain 1: Opinion of PASEO

##### Overall acceptability - Improved contact with HIV care system

When asked about their perception of PASEO, HIV program personnel similarly spoke in positive terms and gave concrete examples of how the intervention affected ALWH. One observation made was that PASEO kept ALWH engaged with the health system, filling a “void” within the HIV care system, as this physician articulated:

We’ve heard good things about the project, from participants’ family members, and we’ve also seen that with several of our patients, especially the difficult ones, it’s been possible to keep them connected with the health system. That’s really good for us because there’s not a system within the hospital that does that. There’s a void there, really, and [PASEO] has come to fill it. (Participant-7)

A peer counselor felt that PASEO helped ALWH manage their care independently, saying, “[PASEO was] guiding them to be able to do things on their own, because they didn’t even have a clue how to get to the hospitals. And now, they’ve restarted the treatments that they were taking before” (Participant-3).

##### What worked - Personalized accompaniment

As with ALWH participants, HIV personnel also emphasized the benefit of tailoring the level of support to the adolescents’ needs, especially when transitioning from pediatric to adult care:

> …for those who left [pediatric HIV services] to go to the other adult hospitals, [PASEO] has been really important because of the accompaniment provided. I know that each of them was given personalized support, because when you get to the adult hospital, it’s different, right? There is no one there to guide you, to support you, nothing. (Participant-3)

The tailoring aspect, it was felt, “humanized” ALWH, suggesting that PASEO went beyond health aspects to include social and educational support, too: “The most important thing [in PASEO] is that we are seeing the life of a human being—right? —in all its context, not only health, but also the social part, the educational part, [as well]” (Participant-1).

##### Support groups

Likewise, feedback from HIV personnel was overwhelmingly positively about PASEO’s support groups; however, there was one respondent who noted the potential that ALWH could also be exposed to other adolescents with undesirable behaviors: “But on the negative side, [in the social support groups, ALWH] probably have access to all kinds of people and could end up falling into risky behaviors” (Participant-4).

##### What did not work or was lacking - Including parents/caregivers in PASEO

No aspect of the intervention elements was noted as faulty; however, one respondent felt that PASEO could be strengthened by including ALWH’s parents in the intervention:

…I am very concerned about the fact, I believe, that there is no consistent work with parents […], because there are cases that I saw where parents did not really support the child or the children […], and when there were in-person group meetings, I don’t know…I feel that there should have been work with [the parents] because they are the in-home support [for their adolescent], right? (Participant-6)

##### Intervention duration

HIV personnel also believed that PASEO would benefit from a longer duration, as in this recommendation from a physician who felt that PASEO should last years:

> …I think that clearly a year is pretty short, perhaps as long as it takes [is the ideal amount of time]. We could say that in this most critical time in adolescence, maybe, I don’t know, 3 years, or if overdoing it, maybe 5 years. I don’t have the answer, but it seems to me like one year is not enough to be able to guarantee long-term continuity of care. (Participant-9)

However, the feasibility of extending the intervention if it were financed by the HIV program was doubted by another HIV care personnel. Still, even a shorter duration of 6 months was felt to have benefit:

> … for the Ministry of Health, I highly doubt that they would want to do it for a year, and I really doubt that they would suddenly accept a full year. They would probably do fewer months, but really, I think that even six months would be enough, and quite important for the patient. (Participant-2)

##### Specific to DOT

DOT, which was delivered in-person pre-COVID and transitioned to telephone and video delivered DOT (“teledot”), garnered differing opinions. On the one hand, some personnel felt that teledot was a strength and should be considered as a HIV program initiative, apart from PASEO: “…well, what actually caught my attention was teledot. It was a really good strategy that we also should do, apart from this project. Seeing the experience and good results that you’ve had, I think we should do it” (Participant-2).

On the other hand, others emphasized the importance that DOT be temporary, as it was in PASEO, given the chronic nature of HIV:

HIV treatment is for life, so thinking about DOT doesn’t make much sense, except in a very conjunctural situation. But really, treatment and counseling must be the aim, they must be incorporated into the lifestyle of someone living with HIV, and that’s their responsibility. (Participant-9)

##### Regarding COVID-19

When asked about how COVID-19 affected PASEO, HIV personnel spoke more generally about the added stress that the pandemic placed on Peru’s HIV treatment program, including hospital closures, medication stock-outs, and related structural issues. In this sense, respondents felt that PASEO somewhat shielded ALWH from the negative impact of COVID-19 on the HIV care delivery system. One participant expressed, “…during the pandemic, some days they didn’t have access to medication [collection]. But because of the support from this project, they managed to continue taking their medications” (Participant-1).

#### Domain 2 - Post-PASEO, opinions, and recommendations

When asked about the future of PASEO, specific to scaling up, two issues arose: who would implement the intervention and how it would be paid for.

*Doubt that the public health system would implement PASEO:* Concern from HIV personnel regarding the public health system’s ability to implement PASEO was expressed; however, the concern appeared less of a budgetary issue and instead related to implementation capacity of the public health system:

[Implementing PASEO is] going to depend on several factors, but what is clear is that it can’t be [part of the public health system’s] services. It must be an external group that intervenes. In what capacity, we’ll have to see, and we’ll see what the budget will be as well… (Participant-9)

Another HIV care personnel believed that implementation would need to encompass all hospitals that ALWH could transition to:

> … more than anything we’ll have to see the resources, funding, to be able to have staff that do the accompaniment part, because that should be done in practically all the hospitals that care for adolescents and then transition them to adult hospitals. (Participant-3)

Finally, a concern arose regarding the potential for low-quality implementation of PASEO and the potential negative consequences for ALWH:

> …maybe I’d worry that [the public health system] doesn’t know how to do [implement PASEO] properly, and there’s a lot of attrition; the kids could lose confidence pretty quickly. So, if they do it and do it poorly, people are going to leave. They’re going to stop following up and stop taking their pills, stop taking their treatment, you know. And that’s a big risk. (Participant-6)

## Discussion

We found that a novel CBA intervention to support ART adherence among ALWH in Lima, Peru—PASEO—was highly acceptable to both ALWH and HIV personnel. Among adolescents, tailored accompaniment and social support groups were especially favored and characterized as emotionally and psychologically transformative. Notably, even participants that engaged less with the intervention found PASEO valuable. Likewise, HIV personnel spoke positively about PASEO, citing favorable experiences they heard from their adolescent patient-participants.

PASEO aimed to improve unsuppressed viremia among ALWH (33); accordingly, the intervention focused on identifying and resolving ALWH’s ART adherence barriers. However, when ALWH spoke about PASEO, responses invariably centered on how each core element made them *feel* rather than the activity itself. ALWH frequently reported improved self-acceptance as they developed emotional bonds with other ALWH. PASEO’s core components appear aligned with the psychosocial stage of development “identity versus role confusion”, in which adolescents form their self-image and sense of direction in life, and in doing so, benefit from greater self-confidence, independence, and an enhanced ability to authentically relate to others [38].

DOT for HIV has yielded mixed findings among adults [39], but in PASEO, ALWH framed DOT as supportive and caring training for independent ART adherence rather than observed verification of medication ingestion. This finding corroborates a study among Peruvian adults with HIV that reported perceived psychosocial outcomes as the primary benefit of DOT [40]. In that study, the stress buffer theory—which posits that community support mitigates perceptions of threat and improves both clinical and psychosocial outcomes—may explain the process by which PASEO participants benefited from DOT [40, 41]. Further, our findings appear to support the positive impact found in a study of community-based DOT for youth with HIV in which adolescents experienced reduced depression and increased coping skills [42].

No negative aspects of PASEO’s core components were reported; however, one perceived deficit was a lack of interaction with ALWH’s parents and/or caregivers. While PASEO implicitly included ALWH’s parents/caregivers, it may benefit from more explicit involvement in adolescents’ support network, and research supports involving parents to enhance ALWH’s ART adherence [32, 43]. However, we note that even in our small sample of ALWH, over half had lost parent(s) to HIV. Therefore, while PASEO might benefit from the option of a parent/caregiver component, our emphasis on the adolescent appears to have been appropriate.

We were interested to understand differences in perceptions of PASEO between ALWH with early childhood and recent HIV. Practically speaking, most ALWH in Peru with behaviorally acquired HIV will also identify as a sexual and/or gender minority (SGM) [44-46] and may experience issues distinct from non-SGM and/or ALWH living with HIV from birth [47]. While there was the suggestion that separate groups for vertically vs. behaviorally (in this case, recently) acquired HIV could help to share common experiences more easily, overall, ALWH were grateful to hear experiences different than their own and endorsed the mixed-group approach. None of the SGM reported feeling marginalized, information which could have emerged during the individual interviews. This finding might be explained by the balance between individual CBA tailored to each ALWH’s specific needs and the support groups which focused on themes that most ALWH could relate to regardless of their background. From an implementation perspective, forming groups with open membership may be easier than multiple, membership-specific groups in certain settings; further research should explore the pros and cons to both approaches.

If PASEO is found to be efficacious in optimizing ART adherence among ALWH, its impact will be constrained unless it can be adequately scaled. HIV personnel voiced concerns regarding financing the intervention and had discrepant views on its duration. These findings speak to future research steps for PASEO, including economic analysis but also enhanced calibration of the dose of PASEO needed to achieve optimal outcomes.

As a pilot study, our findings are limited to the experiences of the study participants and cannot be generalized to all Peruvian ALWH. Though great effort was made to recruit a highly diverse group of adolescents, the sample lacked broad representation of transgender individuals. Future iterations of PASEO and the ability to extend its generalizability will benefit from greater participation by SGM.

## Conclusions

A multicomponent CBA intervention which addresses physical, mental, reproductive, and psychosocial wellbeing to support ART adherence among ALWH in Peru was highly acceptable. Future research should determine the efficacy and economic impact of the intervention.

## Supporting information

Supplemental File COREQ Checklist

## Data Availability

All data produced in the present study are available upon reasonable request to the authors.

## Authors’ contributions

MFF conceived the study. SS, LL, MW, KR, AR, LS, JTG, HS designed the intervention, which was coordinated by MW and implemented by AR, LS, RE, AL, and HS. LK, EM, ES, and CB provided feedback on intervention design and recruitment. JTG led the qualitative analysis, MW collected qualitative data, which was analyzed by JTG, MW and BN. JTG, MW, BN and MFF drafted the manuscript which was reviewed, edited, and approved by all authors.

## Acknowledgements

Our gratitude is extended to the adolescents that participated in PASEO, with special thanks to the Socios En Salud Youth Advisory Board members who provided critical feedback on the study. Karah Greene is thanked for technical support on manuscript preparation.

## Additional Files

Additional File 1: COREQ checklist

## References

1. United Nations Children’s Fund. UNICEF. HIV and AIDS in adolescents. July 2021. Available from: https://data.unicef.org/topic/adolescents/hiv-aids/

2. World Health Organization. HIV and Youth. Available from: https://www.who.int/maternal_child_adolescent/topics/adolescence/hiv/en/

3. World Health Organization. Health for the world’s adolescents: A second chance in the second decade. Available from: https://apps.who.int/adolescent/second-decade/

4. Slogrove AL, Mahy M, Armstrong A, Davies M-A. Living and dying to be counted: What we know about the epidemiology of the global adolescent HIV epidemic. J Int AIDS Soc. 2017;20(0):21520.

5. Buchholz B, Niehues T. Lost in transition: A risk factor for mortality in youth living with HIV. Pediatr Infect Dis J. 2020;39(2):e14–e6.

6. Tsondai PR, Sohn AH, Phiri S, Sikombe K, Sawry S, Chimbetete C, et al. Characterizing the double-sided cascade of care for adolescents living with HIV transitioning to adulthood across Southern Africa. J Int AIDS Soc. 2020;23(1):e25447.

7. Ritchwood TD, Malo V, Jones C, Metzger IW, Atujuna M, Marcus R, et al. Healthcare retention and clinical outcomes among adolescents living with HIV after transition from pediatric to adult care: a systematic review. BMC Public Health. 2020;20(1):1195.

8. Kapogiannis BG, Koenig LJ, Xu J, Mayer KH, Loeb J, Greenberg L, et al. The HIV continuum of care for adolescents and young adults attending 13 urban US HIV care centers of the NICHD-ATN-CDC-HRSA SMILE collaborative. J Acquir Immune Defic Syndr. 2020;84(1):92–100.

9. Berzosa Sanchez A, Jimenez De Ory S, Frick MA, Menasalvas Ruiz AI, Couceiro JA, Mellado MJ, et al. Mortality in perinatally HIV-infected adolescents after transition to adult care in Spain. Pediatr Infect Dis J. 2021;40(4):347–50.

10. Narla NP, Ratner L, Bastos FV, Owusu SA, Osei-Bonsu A, Russ CM. Paediatric to adult healthcare transition in resource-limited settings: a narrative review. BMJ Paediatr Open. 2021;5(1):e001059.

11. Mbalinda SN, Bakeera-Kitaka S, Lusota DA, Musoke P, Nyashanu M, Kaye DK. Transition to adult care: Exploring factors associated with transition readiness among adolescents and young people in adolescent ART clinics in Uganda. PLoS One. 2021;16(4):e0249971.

12. Laurenzi CA, du Toit S, Ameyan W, Melendez-Torres GJ, Kara T, Brand A, et al. Psychosocial interventions for improving engagement in care and health and behavioural outcomes for adolescents and young people living with HIV: a systematic review and meta-analysis. J Int AIDS Soc. 2021;24(8):e25741.

13. Harris LR, Hoffman HJ, Griffith CJ, Lee N, Koay WLA, Rakhmanina NY. Factors associated with transition of HIV care readiness among adolescents and youth from a specialty pediatric HIV clinic in the United States. AIDS Patient Care STDS. 2021;35(12):495–502.

14. Abaka P, Nutor JJ. Transitioning from pediatric to adult care and the HIV care continuum in Ghana: a retrospective study. BMC health services research. 2021;21(1):462.

15. Mbalinda SN, Bakeera-Kitaka S, Lusota DA, Magongo EN, Musoke P, Kaye DK. Barriers and facilitators for transitioning of young people from adolescent clinics to adult ART clinics in Uganda: unintended consequences of successful adolescent ART clinics. BMC health services research. 2020;20(1):835.

16. Straub DM, Tanner AE. Health-care transition from adolescent to adult services for young people with HIV. Lancet Child Adolesc Health. 2018;2(3):214–22.

17. Kim SH, Gerver SM, Fidler S, Ward H. Adherence to antiretroviral therapy in adolescents living with HIV. J Acquir Immune Defic Syndr. 2014;28(13):1945–56.

18. Ridgeway K, Dulli LS, Murray KR, Silverstein H, Dal Santo L, Olsen P, et al. Interventions to improve antiretroviral therapy adherence among adolescents in low-and middle-income countries: A systematic review of the literature. PLoS ONE. 2018;13(1):e0189770.

19. Foster C, Ayers S, Fidler S. Antiretroviral adherence for adolescents growing up with HIV: understanding real life, drug delivery and forgiveness. Ther Adv Infect Dis. 2020;7:204993612092017.

20. Casale M, Carlqvist A, Cluver L. Recent interventions to improve retention in HIV care and adherence to antiretroviral treatment among adolescents and youth: A systematic review. AIDS Patient Care STDs. 2019;33(6):237–52.

21. Hussen SA, Chakraborty R, Camacho-Gonzalez A, Njiemoun B, Grossniklaus E, Goodstein E, et al. Beyond “purposeful and planned”: varied trajectories of healthcare transition from pediatric to adult-oriented care among youth living with HIV. AIDS Care. 2019;31(1):45–7.

22. Foster C, Fidler S. Optimizing HIV transition services for young adults. Curr Opin Infect Dis. 2018;31(1):33–8.

23. Lanyon C, Seeley J, Namukwaya S, Musiime V, Paparini S, Nakyambadde H, et al. “Because we all have to grow up”: supporting adolescents in Uganda to develop core competencies to transition towards managing their HIV more independently. J Int AIDS Soc. 2020;23 Suppl 5:e25552.

24. Abrams EA, Burke VM, Merrill KG, Frimpong C, Miti S, Mwansa JK, et al. “Adolescents do not only require ARVs and adherence counseling”: A qualitative investigation of health care provider experiences with an HIV youth peer mentoring program in Ndola, Zambia. PLoS One. 2021;16(6):e0252349.

25. Bailey H, Cruz MLS, Songtaweesin WN, Puthanakit T. Adolescents with HIV and transition to adult care in the Caribbean, Central America and South America, Eastern Europe and Asia and Pacific regions. J Int AIDS Soc. 2017;20(Suppl 3):21475.

26. Wouters E, Van Damme W, Van Rensburg D, Masquillier C, Meulemans H. Impact of community-based support services on antiretroviral treatment programme delivery and outcomes in resource-limited countries: a synthetic review. BMC health services research. 2012;12(1):194.

27. Munoz M, Finnegan K, Zeladita J, Caldas A, Sanchez E, Callacna M, et al. Community-based DOT-HAART accompaniment in an urban resource-poor setting. AIDS Behav. 2010;14(3):721–30.

28. Gupta N, Munyaburanga C, Mutagoma M, Niyigena JW, Kayigamba F, Franke MF, et al. Community-based accompaniment mitigates predictors of negative outcomes for adults on antiretroviral therapy in rural Rwanda. AIDS Behav. 2016;20(5):1009–16.

29. Franke MF, Kaigamba F, Socci AR, Hakizamungu M, Patel A, Bagiruwigize E, et al. Improved retention associated with community-based accompaniment for antiretroviral therapy delivery in rural Rwanda. Clin Infect Dis. 2013;56(9):1319–26.

30. Thomson DR, Rich ML, Kaigamba F, Socci AR, Hakizamungu M, Bagiruwigize E, et al. Community-based accompaniment and psychosocial health outcomes in HIV-infected adults in Rwanda: a prospective study. AIDS and behavior. 2014;18(2):368–80.

31. United Nations Children’s Fund. Atlas of adolescents and HIV in Latin America and the Caribbean. Adolescents Country Profiles: Peru 2014.

32. Galea JT, Wong M, Munoz M, Valle E, Leon SR, Diaz Perez D, et al. Barriers and facilitators to antiretroviral therapy adherence among Peruvian adolescents living with HIV: A qualitative study. PLoS One. 2018;13(2):e0192791.

33. Vargas V, Wong M, Rodriguez CA, Sanchez H, Galea J, Ramos A, et al. Community-based accompaniment for adolescents transitioning to adult HIV care in urban Peru: a pilot study. medRxiv. 2021:2021.08.25.21261815.

34. Errea RA, Wong M, Senador L, Ramos A, Ramos K, Galea JT, et al. Impact of SARS-CoV-2 pandemic on adolescents living with HIV in Lima, Peru. Rev Peru Med Exp Salud Publica. 2021;38(1):153–8.

35. Bowen DJ, Kreuter M, Spring B, Cofta-Woerpel L, Linnan L, Weiner D, et al. How we design feasibility studies. Am J Prev Med. 2009;36(5):452–7.

36. SocioCultural Research Consultants, LLC. Dedoose. Version 9.0.17 [software]. Los Angeles, CA; 2021. Available from: https://www.dedoose.com/

37. Goldsmith LJ. Using framework analysis in applied qualitative research. Qual Rep. 2021;26(6):2061–76.

38. Ragelienė T. Links of adolescents identity development and relationship with peers: A systematic literature review. J Can Acad Child Adolesc Psychiatry. 2016;25(2):97–105.

39. Gross R, Tierney C, Andrade A, Lalama C, Rosenkranz S, Eshleman SH, et al. Modified directly observed antiretroviral therapy compared with self-administered therapy in treatment-naive HIV-1-infected patients: a randomized trial. Arch Intern Med. 2009;169(13):1224–32.

40. Shin S, Munoz M, Zeladita J, Slavin S, Caldas A, Sanchez E, et al. How does directly observed therapy work? The mechanisms and impact of a comprehensive directly observed therapy intervention of highly active antiretroviral therapy in Peru. Health Soc Care Community. 2011;19(3):261–71.

41. Cohen S, McKay G. Social support, stress and the buffering hypothesis: A theoretical analysis. Handbook of Psychology and Health (Volume IV): Routledge; 2020. p. 253–67.

42. Garvie PA, Flynn PM, Belzer M, Britto P, Hu C, Graham B, et al. Psychological factors, beliefs about medication, and adherence of youth with human immunodeficiency virus in a multisite directly observed therapy pilot study. J Adolesc Health. 2011;48(6):637–40.

43. Naar-King S, Montepiedra G, Garvie P, Kammerer B, Malee K, Sirois PA, et al. Social ecological predictors of longitudinal HIV treatment adherence in youth with perinatally acquired HIV. J Pediatr Psychol. 2013;38(6):664–74.

44. Silva-Santisteban A, Raymond HF, Salazar X, Villayzan J, Leon S, McFarland W, et al. Understanding the HIV/AIDS epidemic in transgender women of Lima, Peru: results from a sero-epidemiologic study using respondent driven sampling. AIDS Behav. 2012;16(4):872–81.

45. Castillo R, Konda KA, Leon SR, Silva-Santisteban A, Salazar X, Klausner JD, et al. HIV and sexually transmitted infection incidence and associated risk factors among high-risk MSM and male-to-female transgender women in Lima, Peru. J Acquir Immune Defic Syndr. 2015;69(5):567–75.

46. Clark JL, Konda KA, Silva-Santisteban A, Peinado J, Lama JR, Kusunoki L, et al. Sampling methodologies for epidemiologic surveillance of men who have sex with men and transgender women in Latin America: an empiric comparison of convenience sampling, time space sampling, and respondent driven sampling. AIDS Behav. 2014;18(12):2338–48.

47. Fields EL, Bogart LM, Thurston IB, Hu CH, Skeer MR, Safren SA, et al. Qualitative comparison of barriers to antiretroviral medication adherence among perinatally and behaviorally HIV-infected youth. Qual Health Res. 2017;27(8):1177–89.

